# Targeting the catecholamine-cytokine axis to prevent SARS-CoV-2 cytokine storm syndrome

**DOI:** 10.1101/2020.04.02.20051565

**Authors:** Maximilian F. Konig, Mike Powell, Verena Staedtke, Ren-Yuan Bai, David L. Thomas, Nicole Fischer, Sakibul Huq, Adham M. Khalafallah, Allison Koenecke, Ruoxuan Xiong, Brett Mensh, Nickolas Papadopoulos, Kenneth W. Kinzler, Bert Vogelstein, Joshua T. Vogelstein, Susan Athey, Shibin Zhou, Chetan Bettegowda

**Affiliations:** Ludwig Center, Lustgarten Laboratory, and the Howard Hughes Medical Institute at The Johns Hopkins Kimmel Cancer Center, Baltimore, MD, USA; Division of Rheumatology, The Johns Hopkins University School of Medicine, Baltimore, MD, USA; Department of Biomedical Engineering, Institute of Computational Medicine, The Johns Hopkins University, Baltimore, MD, USA; Department of Neurology, The Johns Hopkins University School of Medicine, Baltimore, MD, USA; Department of Neurosurgery, The Johns Hopkins University School of Medicine, Baltimore, MD, USA; Division of Infectious Diseases, The Johns Hopkins University School of Medicine, Baltimore, MD, USA; The Johns Hopkins University School of Medicine, Baltimore, MD, USA; Institute for Computational & Mathematical Engineering, Stanford University, Stanford, CA, USA; Janelia Research Campus, Howard Hughes Medical Institute, Ashburn, VA, USA and Optimize Science; Stanford Graduate School of Business, Stanford University, Stanford, CA, USA

## Abstract

In Coronavirus disease 2019 (COVID-19), the initial viral-replication phase is often followed by a hyperinflammatory reaction in the lungs and other organ systems that leads to acute respiratory distress syndrome (ARDS), the need for mechanical ventilation, and death despite maximal supportive care. As no antiviral treatments have yet proven effective, efforts to prevent progression to the severe stages of COVID-19 without inhibiting antiviral immune responses are desperately needed. We have previously demonstrated that a common, inexpensive, and well-tolerated class of drugs called alpha-1 adrenergic receptor (α_1_-AR) antagonists can prevent hyperinflammation (“cytokine storm”) and death in mice. We here present clinical data that supports the use of α_1_-AR antagonists in the prevention of severe complications of pneumonia, ARDS, and COVID-19.

---

Dysregulated host immune responses are drivers of mortality in pneumonia and acute respiratory distress syndrome (ARDS) caused by a wide range of infections. In Coronavirus disease 2019 (COVID-19), severe acute respiratory syndrome coronavirus 2 (SARS-CoV-2) elicits an exuberant local or systemic immune response in the lung and other sites of viral replication, compromising organ function and leading to high morbidity and mortality(1–4).

Emerging evidence suggests that a subset of COVID-19 is characterized by the development of a cytokine storm syndrome (CSS) that resembles cytokine release syndrome (CRS) in chimeric antigen receptor (CAR)-T cell therapy(2, 4, 5). Hyperinflammation in COVID-19 is associated with elevation of pro-inflammatory cytokines including interleukin (IL)-6, IL-2R, IL-8, tumor necrosis factor-α, and granulocyte-colony stimulating factor(4, 6), similar to the exuberant cytokine production by lung-infiltrating monocytes/macrophages and pneumocytes observed in SARS-CoV-1 and MERS-CoV infection(7). Alveolar inflammation and diffuse alveolar damage impair the infected lungs’ local ability to participate in gas exchange, culminating in ARDS and necessitating mechanical ventilation(8). ARDS is the main driver of mortality of COVID-19. Thus, preventing the hyperinflammation in COVID-19 is critical for avoiding this progression.

One potential target is the IL-6 signaling pathway. IL-6 levels diverge profoundly between survivors and non-survivors in the third week after symptom onset and are predictors of COVID-19 severity and in-hospital mortality(1, 6, 9). Tocilizumab, a monoclonal antibody targeting the IL-6 receptor, is currently being investigated for the treatment of patients with COVID-19-CSS(10). Pending data from randomized controlled trials, retrospective data from 21 patients with severe or critical COVID-19 treated with tocilizumab suggests that inhibition of the IL-6 signaling axis is highly effective(11). However, given the cost, immunosuppression, and potential adverse reactions of tocilizumab, this strategy will likely be restricted to select patients in developed countries.

We have recently shown that CRS observed with bacterial infections, CAR-T cells, and other T cell-activating therapies is accompanied by a surge in catecholamines(12). Catecholamines enhance inflammatory injury by augmenting the production of IL-6 and other cytokines through a self-amplifying feed-forward loop in immune cells that requires alpha-1 adrenergic receptor (α_1_-AR) signaling(12). Prophylactic inhibition of catecholamine synthesis with metyrosine, a tyrosine hydroxylase antagonist, reduced levels of catecholamines and cytokine responses and resulted in markedly increased survival following various inflammatory stimuli in mice. Similar protection against a hyperinflammatory stimulus was observed with the well-tolerated α_1_-AR antagonist prazosin (but not with beta-adrenergic receptor [β-AR] antagonists), demonstrating that this class of drugs can also prevent cytokine storm(12).

To date, no controlled trials have examined the potential benefits of α_1_-AR antagonism for the prevention of CSS and mitigation of ARDS in human subjects. To investigate a role for α_1_-AR antagonists in preventing poor outcomes resulting from pulmonary hyperinflammatory responses, we conducted a retrospective analysis of two cohorts of 45-64 year-old hospitalized patients from the MarketScan Research Database (2007-2015). Some of these patients would be expected to be taking α_1_-AR antagonists for the treatment of chronic conditions unrelated to ARDS such as benign prostatic hyperplasia or hypertension. Due to the difficulty of determining from claims data whether individuals were taking their prescribed medication at any given time, we defined prior use as patients having filled α_1_-AR antagonist prescription (doxazosin, prazosin, silodosin, terazosin, or tamsulosin) in the year preceding the event for more than an aggregate of 180 days. Logistic regression models were used to estimate odds ratios (OR), adjusted odds ratios (AOR), and confidence intervals (CI) correlating receipt of α_1_-AR antagonists with two separate outcome measures: a) progression to requiring invasive mechanical ventilation while in the hospital and b) further progression to death while ventilated. Models were adjusted for comorbid hypertension, ischemic heart disease, acute myocardial infarction, heart failure, chronic obstructive pulmonary disease, diabetes mellitus, and post-traumatic stress disorder identified from health care encounters in the prior year as well as age and year.

The first cohort consisted of patients identified with International Classification of Diseases (ICD)- 9 code 518.82 (which encompasses acute respiratory failure including ARDS). Of the 13,125 men in this cohort, we found 655 patients (5.0%) with prior use of α_1_-AR antagonists. Overall, 15.9% of all patients received invasive mechanical ventilation and 8.2% both were ventilated and died in the hospital. We found that patients with prior use of α_1_-AR antagonists had ∼22% lower incidence of invasive mechanical ventilation compared to non-users (OR=0.75, 95% CI 0.59-0.94, p=0.015; AOR=0.75, 95% CI 0.59-0.95, p=0.019) (Figure 1 B,C). Perhaps more importantly, those patients had a ∼36% lower incidence of both being ventilated and dying in the hospital (OR=0.63, 95% CI 0.37-1.01, p=0.074; AOR=0.59, 95% CI 0.34-0.95, p=0.042) (Figure 1 B,D). By contrast, prior use of beta-adrenergic receptor (β-AR) antagonists was not correlated with either outcome in this cohort (Figure 1 C,D).

**Figure 1.**
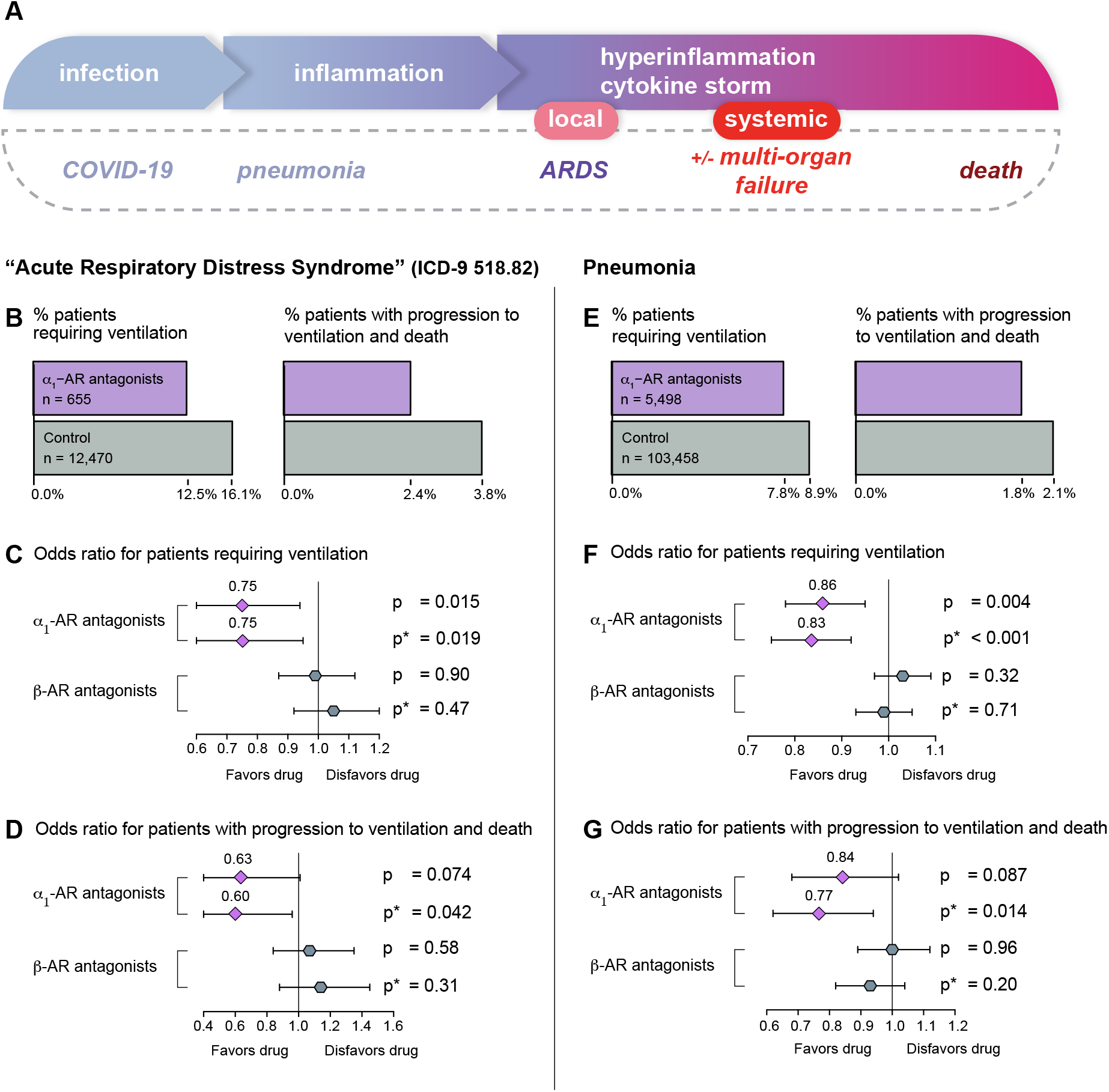
(A) A model of the clinical progression of COVID-19 from local infection to systemic hyperinflammation (“cytokine storm”). The timing and relation of hyperinflammation to specific organ manifestations of severe COVID-19 is an area of uncertainty and investigation. (B-D) Patients from MarketScan Research Database identified by ICD-9 code 518.82 (approximating the diagnosis of ARDS). (B)Number and proportion of patients requiring ventilation (left) or progressing to death (right) with vs without prior use of an α_1_-AR antagonist. (C, D) Forest plots showing odds ratios and 95% confidence intervals (error bars) of progression to invasive mechanical ventilation (C) or progression to death while ventilated (D) with use of α_1_-AR antagonists and β-AR antagonists (control). (E-G) Same as (B-D) but for patients identified with pneumonia (AHRQ category code). The results from both (B-D) and (E-G) are qualitatively similar: α_1_-AR antagonist users have a significantly reduced likelihood of mechanical ventilation and death, whereas β-AR antagonists have no meaningful impact. p and p* correspond to p-values for the unadjusted and adjusted models, respectively.

The second cohort consisted of patients admitted with pneumonia, identified by the Agency for Healthcare Research and Quality’s (AHRQ) pneumonia category, which comprises a number of codes from the ICD-9 and ICD-10, respectively. Of the 108,956 subjects in this cohort, 5,498 patients (5.0%) were taking α_1_-AR antagonist. Overall, 8.9% of all patients received invasive mechanical ventilation and 2.1% both were ventilated and died in the hospital. We found that patients with prior use of α_1_-AR antagonists had ∼13% lower incidence of invasive mechanical ventilation compared to non-users (OR=0.86, 95% CI 0.78-0.95, p=0.004; AOR=0.83, 95% CI 0.75-0.92, p<0.001) (Figure 1 E,F). Further, those patients had a ∼16% lower incidence of both being ventilated and dying in the hospital (OR=0.84, 95% CI 0.68-1.02, p=0.087; AOR=0.77, 95% CI 0.62-0.94, p=0.014) (Figure 1 E,G). By contrast, prior use of β-AR antagonists was not correlated with either outcome in this cohort, with or without adjusting (Figure 1 F,G). All stated results were robust to multiple propensity weighting approaches, including causal forest variants(13). These findings suggest that α_1_-AR antagonists may protect from immune-mediated morbidity and mortality resulting from common lung injury and infection.

Taken together, these results extend preclinical findings to support a clinical rationale for studying α_1_-AR antagonists in the prophylaxis of severe COVID-19 and states of local and systemic immune dysregulation. Prazosin is inexpensive and safe, as has been documented by long-term treatment of millions of patients with benign prostatic hyperplasia, hypertension, and other conditions. However, all drugs can have unanticipated side effects in different clinical contexts, and the incompletely understood relationship between hypertension and COVID-19 suggests caution in using any agent that impacts blood pressure(14).

Given the limitations of retrospective studies (such as this one), prospective clinical trials of α_1_-AR antagonists in high-risk patients will therefore be required to assess their utility in preventing COVID-19-CSS. We emphasize that the extensive experience with using prazosin for other indications should prioritize — *not obviate —* rigorous, controlled clinical research rather than indiscriminate off-label use in patients exposed to or infected with SARS-CoV-2. Such trials could be expeditiously implemented in areas suffering from high infection rates that are overwhelming hospital capacity. To that end, we are actively pursuing clinical trials at multiple institutions and will make our protocols available on http://clinicaltrials.gov/ when approved by the Johns Hopkins Internal Review Board. We encourage readers to contact us and/or launch trials based on these or other compelling retrospective analyses coupled with pathophysiological mechanistic explanations.

## Data Availability

NA

## Acknowledgements

We thank Adam Sacarny (Columbia University) for advice in processing and analyzing health care claims data. Dr. Sacarny was not compensated for his assistance. This study used the IBM MarketScan Research Databases. Research including data analysis has been partially supported by funding from Microsoft Research.

## Disclosure

In 2017, The Johns Hopkins University (JHU) filed a patent application on the use of various drugs to prevent cytokine release syndromes, on which V.S., R.B., N.P., B.V., K.W.K., and S.Z. are listed as inventors. JHU will not assert patent rights from this filing for treatment related to COVID-19.

## Notes

### Competing Interest Statement

Drs. Staedtke, Bai, Papadopoulos, Kinzler, Vogelstein, and Zhou are inventors on a patent application filed by Johns Hopkins University entitled “Preventing cytokine release syndrome”. Biomed Valley Discoveries has previously funded the work described in Staedke et al. and has licensed related patents from Johns Hopkins University. Drs. Papadopoulos, Kinzler, Vogelstein, and Zhou are entitled to a share of royalties received by the University from Biomed Valley Discoveries. The terms of these arrangements are managed by Johns Hopkins University in accordance with its conflict of interest policies. Dr. Bettegowda reports personal fees from Depuy-Synthes and Bionaut Pharmaceuticals outside the submitted work.

### Funding Statement

Dr. Konig was supported by the National Institute of Arthritis and Musculoskeletal and Skin Diseases of the National Institutes of Health under Award no. T32AR048522. This work was further supported by The Virginia and D.K. Ludwig Fund for Cancer Research, The Lustgarten Foundation for Pancreatic Cancer Research, and the BKI Cancer Genetics and Genomics Research Program.

